# Remote monitored physiological response to therapeutic escalation and clinical worsening in patients with pulmonary arterial hypertension

**DOI:** 10.1101/2023.04.27.23289153

**Authors:** Jennifer T Middleton, Sarah Binmahfooz, Hamza Zafar, Junaid Patel, Cameron Ashraf, Jake, Dharshan Neelam-Naganathan, Christian Battersby, Charlotte Pearson, Chloe Roddis, Stefan Roman, Jenna Ablott, Ashwin Reddy, Lisa Watson, Jennifer Dick, Andreas Kyriacou, Paul D Morris, Frances Varian, Neil Hamilton, Iain Armstrong, Judith Hurdman, Abdul Hameed, Athanasios Charalampopoulos, Theophile Bigirumurame, Shaun K. W. Hiu, James M. S. Wason, Andrew J Swift, A A Roger Thompson, Robin Condliffe, Charlie Elliot, David G Kiely, Mark Toshner, Alexander M K Rothman, the United Kingdom Pulmonary Hypertension (UNIPHY) Clinical Trials Network and the National Cohort Study of Idiopathic and Heritable PAH

## Abstract

**Background:** International guidelines recommend regular, hospital-based risk stratification to aid assessment and management of patients with pulmonary arterial hypertension. Technological advances enable daily, remote measurement of cardiopulmonary physiology and physical activity that have the potential to provide early evaluation of therapeutic efficacy and facilitate early intervention based on the physiological changes that precede clinical events. We sought to investigate the relationship between remote-monitored parameters and the COMPERA 2.0 4-strata risk score and evaluate physiological changes following therapeutic escalation and prior to clinical worsening events.

**Methods:** Eighty-seven patients with pulmonary arterial hypertension were implanted with insertable cardiac monitors including a nested set of twenty-eight patients also implanted with a pulmonary artery pressure monitor. Hospital measured and remote monitored physiological parameters were evaluated by 4-strata COMPERA 2.0 risk score. A time stratified bidirectional case-crossover study was undertaken to evaluate physiological changes at the time of therapy escalation and clinical worsening events in the nested group with insertable cardiac and pulmonary artery pressure monitors. A summary measure of remote physiological risk was calculated as the sum of the z-score of physical activity, heart rate reserve and total pulmonary resistance and applied to remote monitoring data.

**Results:** Insertable cardiac monitor-measured physical activity, heart rate variability and heart rate reserve were decreased and night heart rate increased in patients with increasing COMPERA 2.0 score (p<0.0001). Daily physical activity was related to incremental shuttle walk distance (p<0.0001) but not six-minute walk distance. Following therapeutic escalation mean pulmonary artery pressure and total pulmonary resistance were reduced and cardiac output, and physical activity increased at 7, 4, 22, and 42 days, respectively (p<0.05). Clinical worsening events were preceded by increased mean pulmonary artery pressure and total pulmonary resistance, reduced cardiac output and physical activity (p<0.05). Applying a remote physiological risk score to remote-monitored data demonstrated that following a clinically indicated increase in therapy, a reduction in physiological risk was identifiable at day three, and preceding a clinical worsening event, an increase in adverse physiology was observable at day - 16.

**Conclusion:** Approved devices accurately identify change in physiology in patients with pulmonary arterial hypertension following therapeutic intensification and before clinical worsening. A remote assessment of haemodynamic and cardiac monitoring may facilitate personalised, proactive medicine and innovative clinical study designs.

**Condensed Abstract:** Technological advances provide the capacity to remotely measure cardiopulmonary physiology. In 87 patients with insertable cardiac monitors and a nested group 28 patients with pulmonary arterial hypertension implanted with pulmonary artery pressure monitors, significant improvements in cardiopulmonary function and physical activity were observed following therapeutic escalation and preceding clinical worsening events. The study highlights the potential of remote monitoring for personalised management, early therapeutic evaluation, and innovative clinical trial designs in patients with pulmonary hypertension.

**Twitter (X) post:** #PHPEEPS Remote monitoring shows improved cardiopulmonary function just 7 days after therapy adjustments, and adverse changes 12 days before a worsening event. The future of personalised care?

**Learning points:** Pulmonary artery pressure monitor and insertable cardiac monitors offer safe and reliable data capture of physiological risk markers that change in response to therapy and preceding clinical worsening events.

Remote monitored measures of physiology differ between patients with low, int-low, int-high and high risk of one-year mortality stratified by COMPERA 2.0 4-strata risk model.

Remote risk evaluation may facilitate personalised medicine and proactive management for early evaluation of therapeutic efficacy and detection of clinical worsening.

**Plain Language Summary:** This study was undertaken in 87 patients diagnosed with pulmonary arterial hypertension (PAH). Treatments in PAH are based on a risk classification system with the aim of achieving a low-risk group. The usual in-hospital method uses the COMPERA 2.0 risk model which combines a field walk test, NT-proBNP (blood test) and World Health Organisation Functional Classification (WHO FC) which categorises level of breathlessness during everyday activity. The evidence for this is linked to risk of death, classified into four groups: low, intermediate-low, intermediate-high, and high risk. The aim of this study was to see whether newer medical technologies could grade risk in a remote setting.

The two technologies used in this study are safe and approved for use. The first is a pulmonary artery pressure monitor (CardioMEMS) that measures the pressure in the lungs. It is implanted during right heart catheterisation (RHC). Measurements can be taken at home and sent securely to a medical database for the healthcare team to view. Please see the plain graphical summary figure for more information on the CardioMEMS device. The second technology is an insertable cardiac monitor (ICM), which is implanted under the skin using local anaesthetic, and sends remote readings such as physical activity and heart rate. Both technologies were implanted into a subgroup of patients to investigate whether these technologies could help classify risk from home, and whether they could detect response to new treatments, or signs that a condition may be getting worse. 28 patients with both these devices took part in the study and a further 59 had an ICM only. A remote risk score was calculated using 3 things: physical activity, heart rate reserve (HRR: difference between maximum heart rate for age and resting heart rate) from the ICM and total pulmonary resistance (TPR: a measure of the pressure and flow through the lungs) from the CardioMEMS. The results showed that these measures could classify risk as well as the in-hospital COMPERA 2.0 model. The remote risk score detected response to treatment as early as 6 days and clinical worsening as early as 12 days before an event (e.g. hospitalisation) in the group observed.

**Patient and Public Involvement and Engagement (PPIE):** The study was developed following the 2017 Pulmonary Hypertension Association UK (PHA UK) survey in which 39% of patients reported difficulties attending hospital for appointments.^1^ A subsequent remote monitoring survey (2021) was positively received, with key themes highlighting benefits of ‘improving [disease] understanding’, ‘personalising treatment’, and ‘reducing interruptions or unnecessary visits’.^2^ Patients from the study and volunteers from PHA UK provided feedback on the results of the research. Amendments were made to the lay summary and a graphical summary was introduced following this feedback. There was universal agreement that participation in the study was beneficial to patients and future research. Participants involved in the study agreed the devices offer enhanced accessibility to non-invasive risk stratification and improvements in home-based care with minimal personal effort. Furthermore, the minimally invasive devices offered empowerment, confidence, and reassurance, with “opportunity to play an active role in [their] health and personal wellbeing” and “greater confidence with day-to-day living”. No incentives were offered for the PPIE in this study.

Plain Graphical Summary:
CardioMEMS implantation covering frequently asked questions (FAQ). Created with BioRender.com

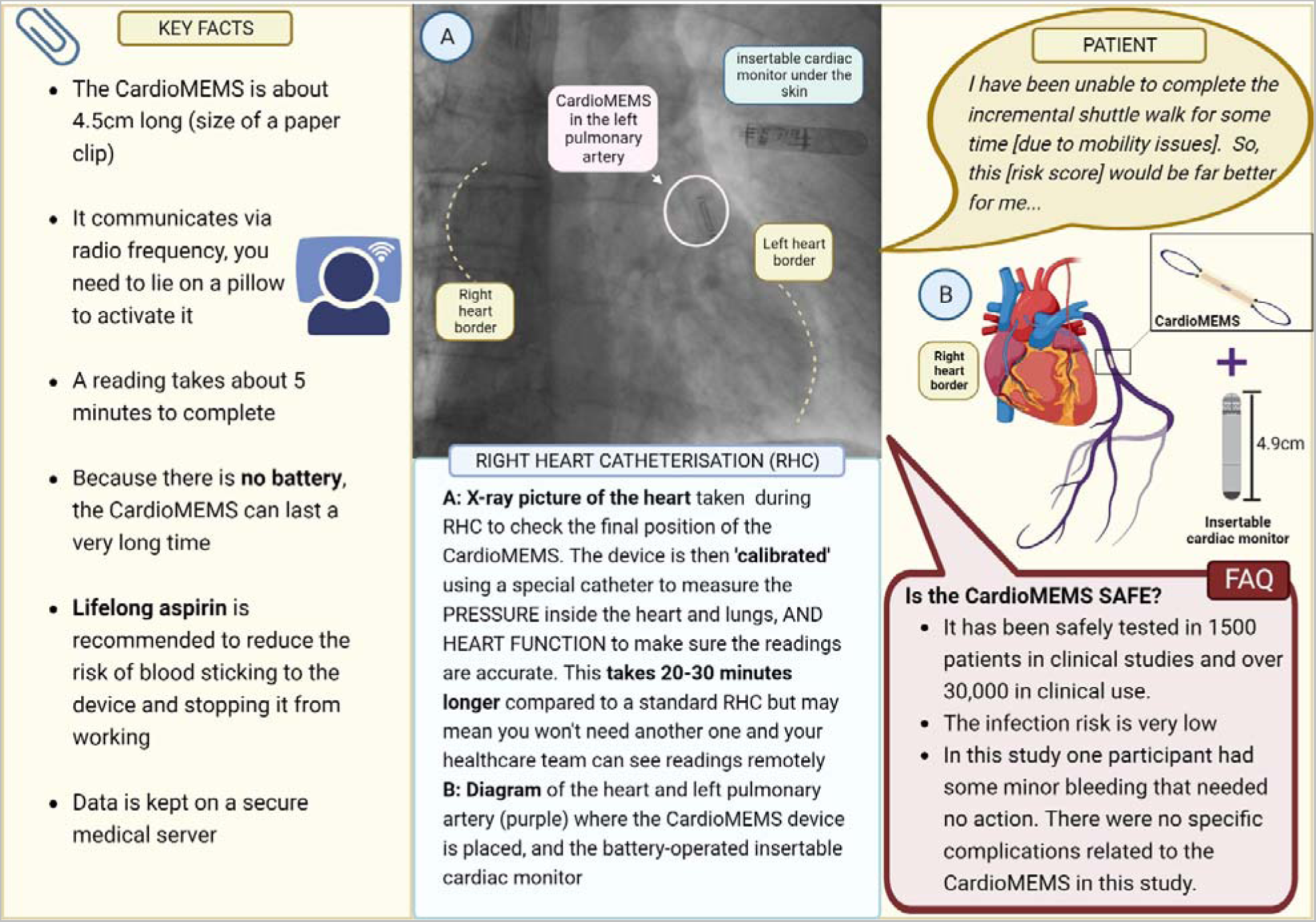

## Introduction

Pulmonary arterial hypertension is a life-shortening condition driven by remodelling and constriction of pulmonary arterioles that leads to increased pulmonary vascular resistance and pulmonary artery pressure, and right heart failure.^3^ Licensed therapies target vasoconstriction and vasodilatation of the pulmonary vasculature^4^ and with treatment 5-year survival is ~70%.^5–7^ However, not all patients respond to therapy,^8^ side effects are common.^9,10^

European guidelines provide an expert-opinion-based risk-stratification score to aid treatment decisions based on hospital-based assessment of symptoms, exercise capacity, and right ventricular function.^11^ Patients who improve to a low-risk profile at follow-up have increased survival compared to those who fail to demonstrate clinical improvement.^12^ Current clinical approaches aim to improve and maintain patients in the low-risk group^13^ and as such, there is significant interest in the identification of remote measures that may offer the potential for frequent, early evaluation of clinical efficacy following treatment change or identify the physiological changes that precede clinical deterioration.

Technological advances have led to the development of minimally invasive devices that measure cardiopulmonary haemodynamics,^14,15^ cardiac physiology^16^ and physical activity,^17^ daily, from the patient’s home. In patients with pulmonary arterial hypertension, we undertook a preliminary study to evaluate the capacity of advanced haemodynamic and cardiac monitoring devices to detect changes in physiology following clinically indicated therapeutic escalation and prior to clinical worsening events.

## Methods

### Advanced haemodynamic and insertable cardiac monitoring

Patients with a confirmed diagnosis of PAH, in WHO functional class (FC) III were enrolled in **F**easibility of Novel Clinical Trial **I**nfrastructure, Design and **T**echnology for Early Phase Studies in Patients with **P**ulmonary **H**ypertension (FIT-PH, 19/YH/0354) and the UK National Cohort Study of Idiopathic and Heritable Pulmonary Arterial Hypertension (13/EE/0203). A pulmonary artery pressure monitor (CardioMems, Abbott) and insertable cardiac monitor (LinQ, Medtronic) were implanted using standard techniques and remote monitoring data collected via regulatory approved online portals.

### Devices and data handling

Cardiac output was calculated using a proprietary algorithm (Abbott) based on the pulmonary artery pressure waveform, mean pulmonary artery pressure, heart rate, and a reference cardiac output measured at implant that has been tested against clinical right heart catheter data, demonstrating non-inferiority to clinically used cardiac output measurement,^18,19^ and utility in patients with pulmonary arterial hypertension.^20^ Physical activity was measured from the single axis accelerometer in the insertable cardiac monitor. A minute is considered active if a threshold is reached incorporating deflection number and magnitude, which has proven responsive in capturing activities of daily living that correlate with clinical events in patients with heart failure and/or atrial fibrillation.^17,21–24^

### Clinical management and events

Assessment of baseline risk was made using the 4-strata COMPERA 2.0 criteria.^11,12^ A multi-professional team (pulmonologist, cardiologist, pharmacist, and specialist nurse) reviewed data, adjudicated events and directed treatment. Disease progression/worsening was defined as disease-related hospitalisation, escalation of disease-specific therapy, or the need for lung transplantation as judged by the physician,^9,10,25^ and as a decrease from baseline of at least 15% in the incremental shuttle walk test accompanied by a worsening in WHO-FC.^9,10,25^

### Waveform evaluation

Suspected artefactual readings were identified by a proprietary automatic algorithm or those two SD from the rolling mean pulmonary artery pressure. Manual review of the pressure waveform was undertaken by two qualified physicians to identify those resulting from non-rested physiological state, post walk-test, ventricular ectopic beats, transmission failure, incorrect frequency detection or damped waveforms (Supplemental Figure 1).

### Study Design

A time-stratified bidirectional case–crossover approach was taken to provide remote monitoring data from the time of clinical events and individual patient matched control periods.^26^ Control data comprises randomly selected time periods of a matched duration not overlapping event periods (Supplemental Figure 2).

### Remote risk score

The remote risk score was calculated as ztotal pulmonary resistance −zheart rate reserve −zheart rate variability −zphysical activity and applied to remote monitoring data at the time of clinical.

### Statistical Analysis

Data were tested for normality using the Kolmogorov–Smirnov test and statistical comparisons made using Students’ t-test and ANOVA as appropriate. Statistical analysis was performed using IBM SPSS version 27 and Prism 9 for macOS (version 9.3.0).

## Results

### Study population device implantation

Between 26^th^ of Jan 2020 and 25^th^ of May 2022, 87 patients with pulmonary arterial hypertension were implanted with an insertable cardiac monitor, 28 of whom were implanted with a pulmonary artery pressure monitor (table 1). The mean age was 48.9 years +/−18.1, 21 patients (75%) were women and 25 (89%) were Caucasian. There were no device-related serious adverse events and two device-related adverse events (one minor haemoptysis and one insertable cardiac monitor wound dehiscence at day 3, supplemental table 1). Following implantation, data completeness was 100% for the insertable cardiac monitor and 91.7% for the pulmonary artery pressure monitor (Supplemental Figure 3).

**Table 1:**
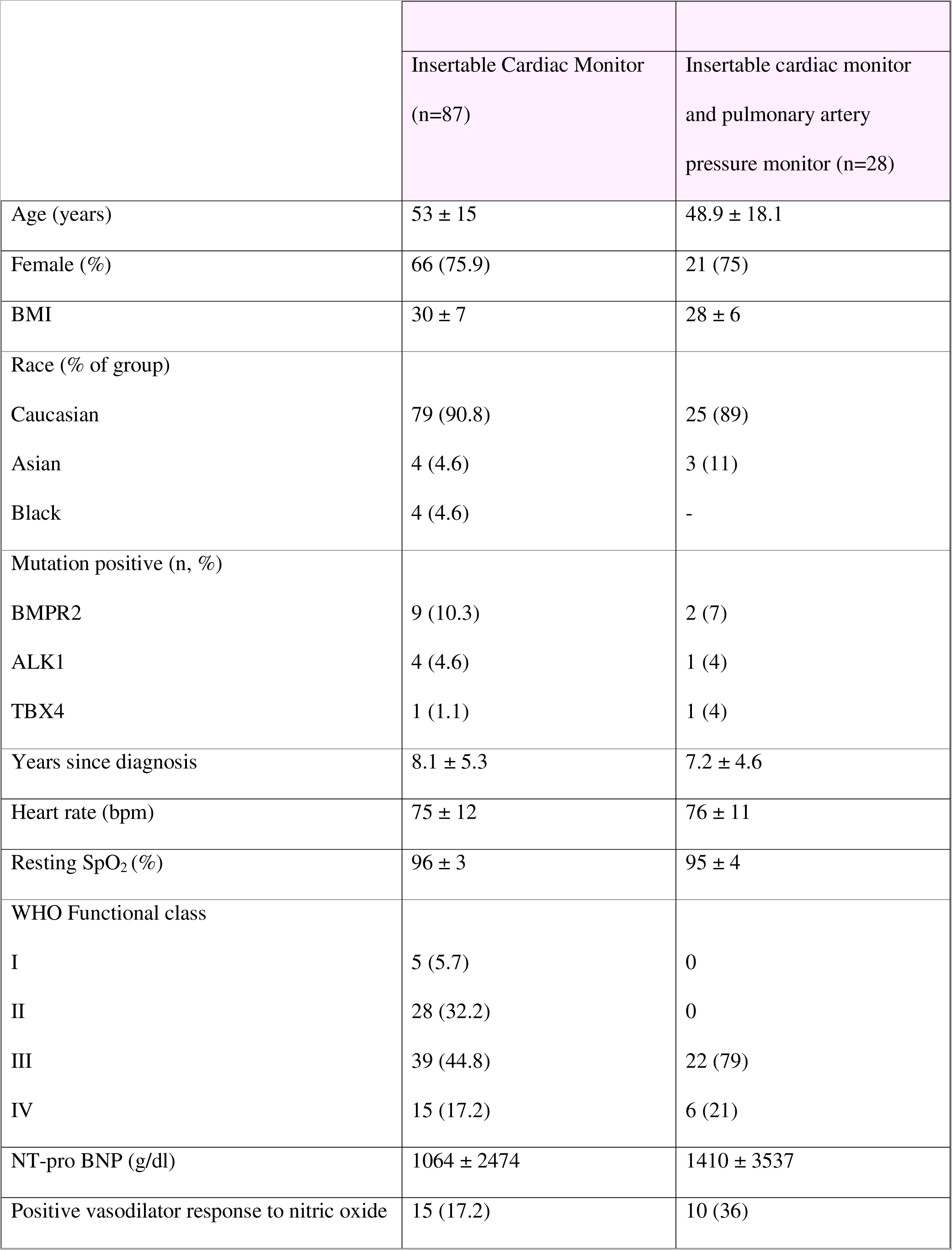
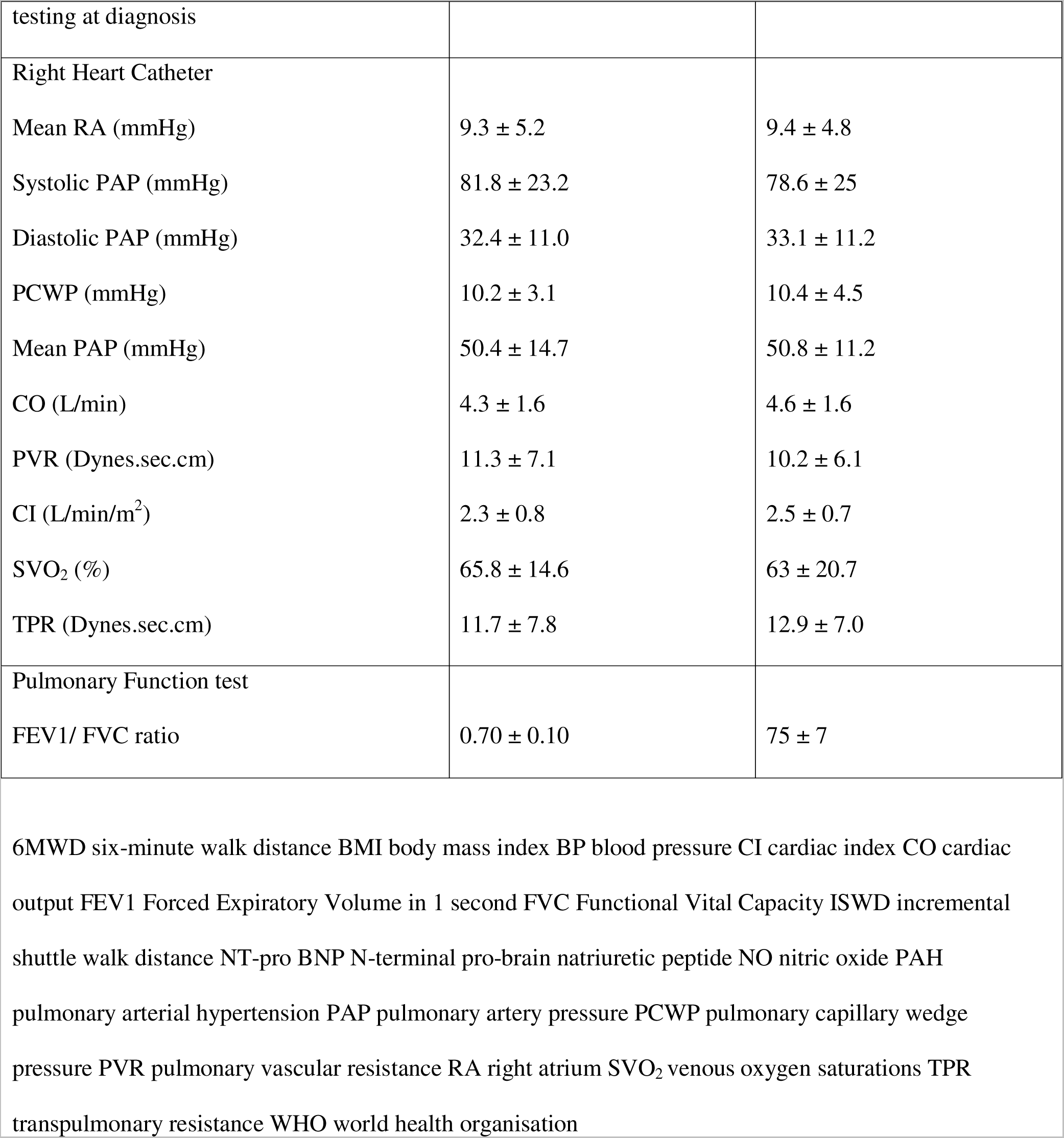
Baseline demographics for patients with an implantable cardiac monitor.

|  | Insertable Cardiac Monitor<br>(n=87) | Insertable cardiac monitor<br>and pulmonary artery<br>pressure monitor (n=28) |
| --- | --- | --- |
| Age (years) | 53 ± 15 | 48.9 ± 18.1 |
| Female (%) | 66 (75.9) | 21 (75) |
| BMI | 30 ± 7 | 28 ± 6 |
| Race (% of group) |  |  |
| Caucasian | 79 (90.8) | 25 (89) |
| Asian | 4 (4.6) | 3 (11) |
| Black | 4 (4.6) | - |
| Mutation positive (n, %) |  |  |
| BMPR2 | 9 (10.3) | 2 (7) |
| ALK1 | 4 (4.6) | 1 (4) |
| TBX4 | 1 (1.1) | 1 (4) |
| Years since diagnosis | 8.1 ± 5.3 | 7.2 ± 4.6 |
| Heart rate (bpm) | 75 ± 12 | 76 ± 11 |
| Resting SpO<sub>2</sub> (%) | 96 ± 3 | 95 ± 4 |
| WHO Functional class |  |  |
| I | 5 (5.7) | 0 |
| II | 28 (32.2) | 0 |
| III | 39 (44.8) | 22 (79) |
| IV | 15 (17.2) | 6 (21) |
| NT-pro BNP (g/dl) | 1064 ± 2474 | 1410 ± 3537 |
| Positive vasodilator response to nitric oxide | 15 (17.2) | 10 (36) |
| testing at diagnosis |  |  |
| Right Heart Catheter |  |  |
| Mean RA (mmHg) | $9.3 \pm 5.2$ | $9.4 \pm 4.8$ |
| Systolic PAP (mmHg) | $81.8 \pm 23.2$ | $78.6 \pm 25$ |
| Diastolic PAP (mmHg) | $32.4 \pm 11.0$ | $33.1 \pm 11.2$ |
| PCWP (mmHg) | $10.2 \pm 3.1$ | $10.4 \pm 4.5$ |
| Mean PAP (mmHg) | $50.4 \pm 14.7$ | $50.8 \pm 11.2$ |
| CO (L/min) | $4.3 \pm 1.6$ | $4.6 \pm 1.6$ |
| PVR (Dynes.sec.cm) | $11.3 \pm 7.1$ | $10.2 \pm 6.1$ |
| CI (L/min/m<sup>2</sup>) | $2.3 \pm 0.8$ | $2.5 \pm 0.7$ |
| SVO<sub>2</sub> (%) | $65.8 \pm 14.6$ | $63 \pm 20.7$ |
| TPR (Dynes.sec.cm) | $11.7 \pm 7.8$ | $12.9 \pm 7.0$ |
| Pulmonary Function test |  |  |
| FEV1/ FVC ratio | $0.70 \pm 0.10$ | $75 \pm 7$ |

### Relationship of baseline haemodynamic measures to 4-strata COMPERA 2.0 risk model

At enrolment, WHO functional class and NT-proBNP were increased with COMPERA 2.0 risk classification group (figure 1A-B), however ECG measured heart rate differentiated only those in intermediate-low risk compared to those at high risk (figure 1C). Similarly, pulmonary artery pressure was increased in the high-risk group compared to low and int-low groups and total pulmonary resistance was increased in the high-risk group compared to low, int-low and int-high groups. Cardiac output did not differ between risk groups (figure 1D-F).

**Figure 1:**
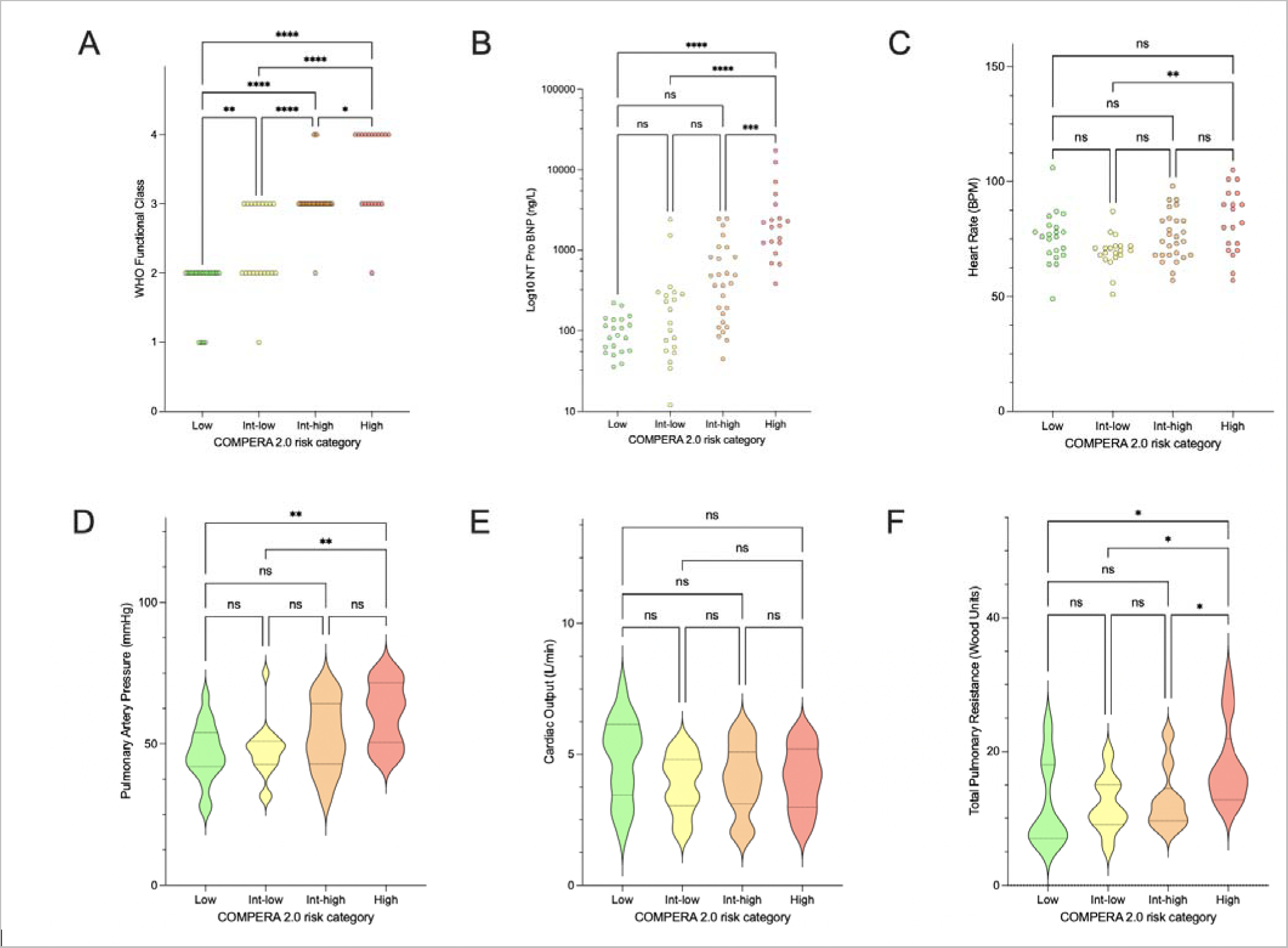
Baseline in-hospital measures from patients with pulmonary arterial hypertension and an insertable cardiac monitor. In-hospital measures of WHO functional class (A), Ntpro-BNP (B) ECG heart rate (C) pulmonary artery pressure (D) cardiac output (E) total pulmonary resistance (F) by 4-strata COMPERA 2.0 score (n=87, two-way ANOVA with Tuckey correction, *p<0.05, **p<0.01, ***p<0.001, ****p<0.0001).

### Relationship of remote monitored cardiac and physical activity measures to 4-strata COMPERA 2.0 risk model

Over the six months following implantation, remote-monitored cardiac measures accurately identified differences in physiology in patient groups stratified by 4-strata COMPERA 2.0 risk model. Night heart rate increased, and heart rate reserve (day heart rate – night heart rate) and heart rate variability reduced with each COMPERA 2.0 risk group (figure 2). A u-shaped relationship was observed between increasing risk and day heart rate (figure 2). Notably a negative bias was observed when comparing ECG-measured heart rate and night heart rate measured by insertable cardiac monitor (figure 2E). Remote monitored physical activity was reduced with increased COMPERA 2.0 risk (Figure 3A). A correlation between physical activity and incremental shuttle walk distance was present, however, no relationship was identified with 6-minute walk distance (figure 3B-C).

**Figure 2:**
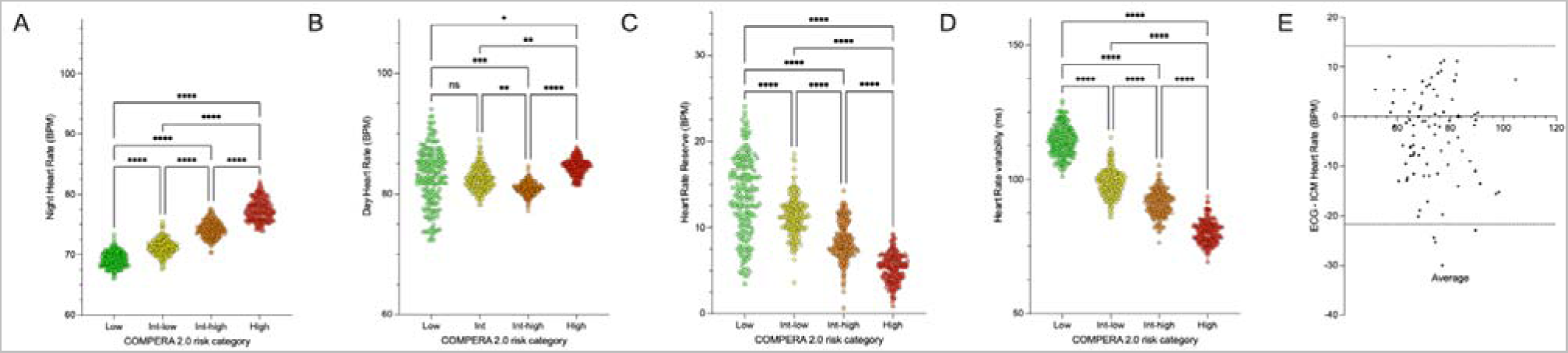
Remote-monitored cardiac measures in patients with pulmonary arterial hypertension and an insertable cardiac monitor. Daily remote measures of night heart rate (A), day heart rate (B), heart rate reserve (C) and heart rate variability (D) over the first 6 months following implantation of insertable cardiac monitors in patients with pulmonary arterial hypertension by 4-strata COMPERA 2.0 score (n=87, two-way ANOVA with Tuckey correction, *p<0.05, **p<0.01, ***p<0.001, ****p<0.0001). Bias of baseline ECG and insertable cardiac monitor measured night heart rate (E).

**Figure 3:**
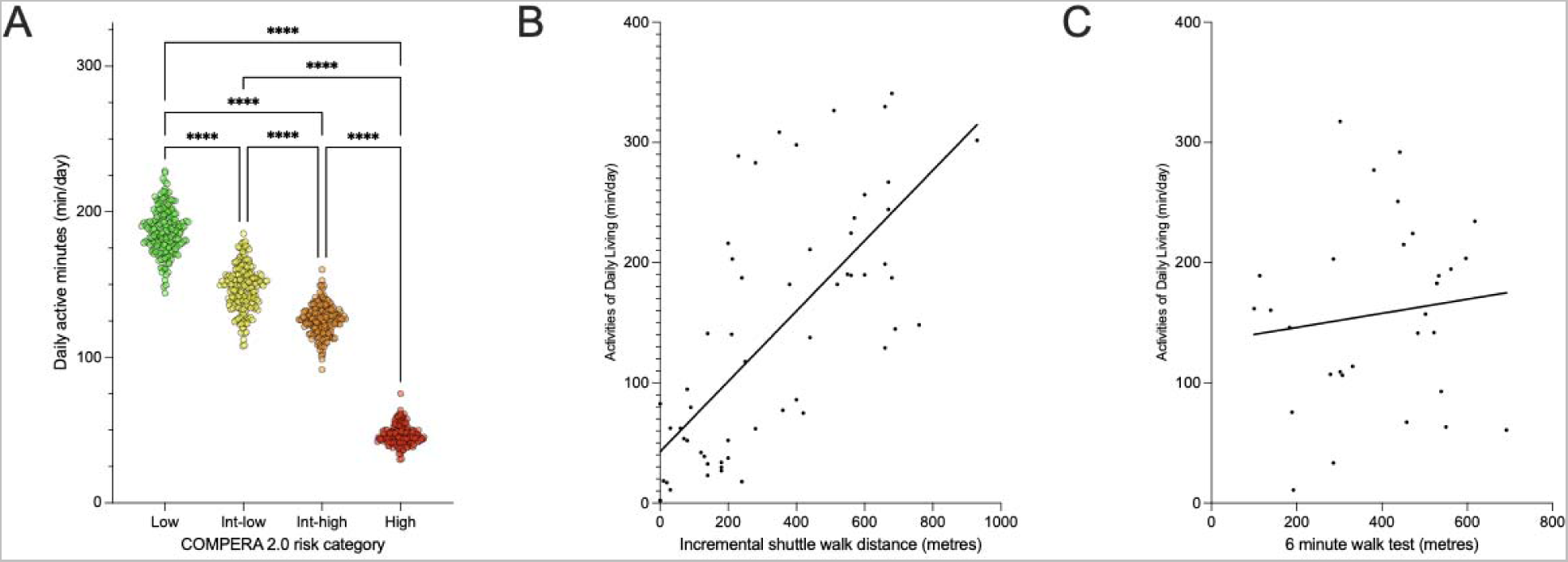
Remote-monitored physical activity in patients with pulmonary arterial hypertension and an insertable cardiac monitor. A: Physical activity over the first 6 months following implantation of insertable cardiac monitors in patients with pulmonary arterial hypertension by 4-strata COMPERA 2.0 score (n=87, two-way ANOVA with Tuckey correction, ****p<0.0001). Relationship of physical activity to baseline incremental shuttle walk distance (F, Pearson, r^2^=0.50, p<0.0001) and 6-minute walk distance (G, Pearson, r^2^=0.02, p=NS).

### Physiological response to therapeutic escalation in patients with pulmonary arterial hypertension

In patients implanted with both an insertable cardiac monitor and pulmonary artery pressure monitor survival was 96% at 6-months and 93% 12-months. Expected survival, estimated using individual patient data input into the French registry equation, was 89% and 83% at 6 and 12 months, respectively.^27^ From implantation to follow-up there was no change in estimated glomerular filtration rate (76.3 +/−13.5 ml/min/1.73 m^2^ vrs 75.2 +/− 12.4 ml/min/1.73 m^2^; p = NS). At enrolment, 7 patients were within 6 weeks of treatment initiation and 21 patients were established on stable therapy. Three patients were receiving oral monotherapy with CCB, 14 patients were receiving dual oral therapy, 11 patients were receiving oral therapy plus inhaled or intravenous prostanoid or IP receptor agonist (table 2).

**Table 2:**
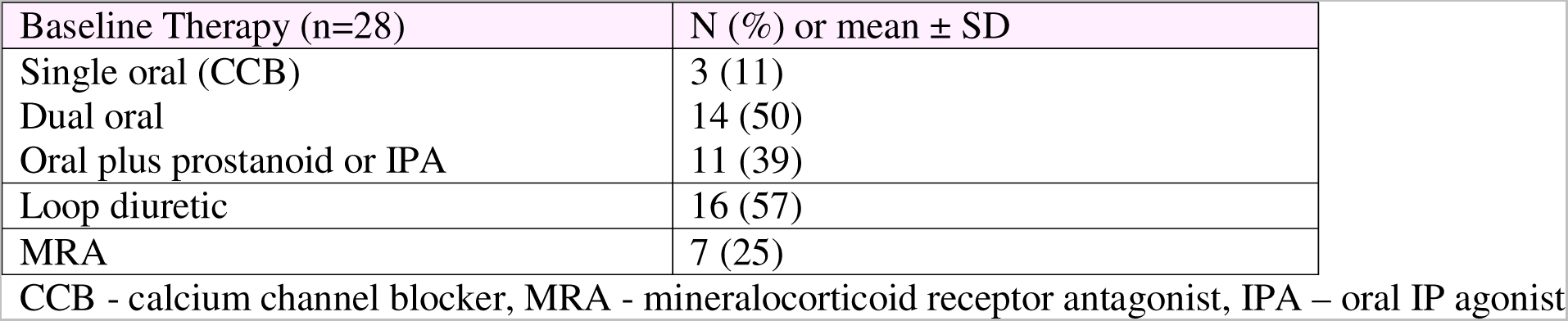
Baseline therapy – FIT-PH.

| Baseline Therapy (n=28) | N (%) or mean ± SD |
| --- | --- |
| Single oral (CCB) | 3 (11) |
| Dual oral | 14 (50) |
| Oral plus prostanoid or IPA | 11 (39) |
| Loop diuretic | 16 (57) |
| MRA | 7 (25) |

2 CCB - calcium channel blocker, MRA - mineralocorticoid receptor antagonist, IPA – oral IP agonist

To evaluate the changes in physiology following a clinically directed therapeutic escalation we examined remote monitoring data in the 30-days preceding, and 60 days following the addition or increase in disease specific therapy. Clinically indicated changes in therapy were: up titration of CCB (3), addition of PDE (3), addition of ERA (3), addition of IP receptor agonist (4) and initiation of parental prostanoid (5). Following clinician-directed initiation or increase in therapy ISWD (183m +/− 49 to 345m +/− 47, p<0.001), WHO functional class (p<0.001) and NT-proBNP (779 pg/ml +/−243 to 402 pg/ml +/−139, p<0.0001), were improved with a mean time to assessment of 4.4 months +/− 0.4 (Figure 4A-D). During periods without a therapeutic escalation or clinical worsening event (assessed by the multi professional team) remote monitored physiological parameters were stable. Consistent with identified improvements in established measures, remote monitored mean pulmonary artery pressure (day 0-30: −6.3, −4.6 to −8.2 mmHg, p<0.0001; day 30-60: −8.1, −5.1 to −11.2 mmHg, p<0.0001), total pulmonary resistance (day 0-30: −2.5, −1.4 to −3.6 WU, p<0.0001; day 30-60: −3.2, −1.8 to −4.6 WU, p<0.0001) and night heart rate (day 0-30: −7 bmp, −2 to −12 bpm, p<0.05; day 30-60: −9 bpm, −1 to −17 p<0.05) were reduced and cardiac output (day 0-30: 0.4, 0.2-0.6 L/min, p<0.01; day 30-60: 0.7, 0.3-1.1 L/min, p<0.001) and physical activity (day 0-30: 15.2, −10.3-40.7 min/day, p<NS; day 30-60: 31.0, 1.0-62.0 min/day, p<0.05) increased 60 days following treatment change (Figure 4E-J, p<0.05). Daily measures of these parameters taken from the patients home demonstrate that mean pulmonary artery pressure and total pulmonary resistance were reduced, and cardiac output and physical activity increased at days 4, 7, 22 and 42 respectively.

**Figure 4:**
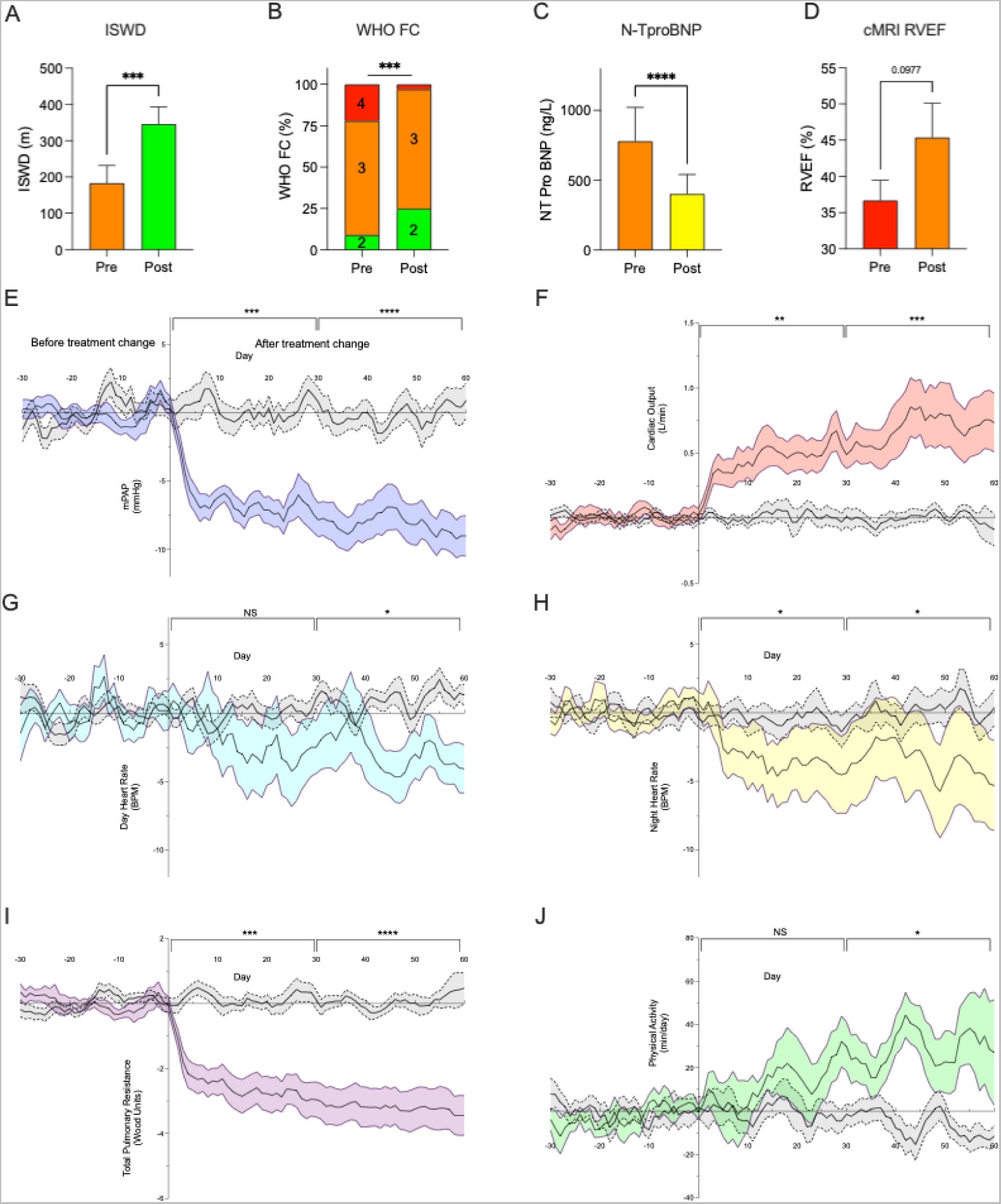
Established risk measures and remote-monitored physiology following clinically indicated treatment change. A. Incremental shuttle walk distance (ISWD), B. WHO functional class (WHO FC), C. NTProBNP and D. right ventricular ejection fraction from cardiac MRI (cMRI-RVEF). Mean +/−SEM, Wilcoxon, ***p<0.001, ****p<0.0001 A-C: n=18, D: n=9). E: mean pulmonary artery pressure, F: cardiac output, G: day heart rate, H: night heart rate; I total pulmonary resistance; J: physical activity. Data is presented with treatment change at day 0 with days –30 to day −1 as days preceding, and days +1 to day +60 as days following treatment change. Control group comprises matched 90-day periods on stable therapy (grey). Treatment change n=18, mean +/−SEM, *p<0.05, p<0.01, ***p<0.001, ****p<0.0001 one-way ANOVA with Dunnett’s correction.

### Physiological response to clinical worsening in patients with pulmonary arterial hypertension

To evaluate the changes related to disease worsening remote monitoring data was evaluated in the 30-days preceding and following clinical worsening events (defined in clinical management and events section of the methods). Following a clinical worsening event: ISWD (343m +/−71 to 241m +/−65, p<0.001), WHO functional class (p<0.001) and NT-proBNP (757 pg/ml +/− 319 to 2472 pg/ml +/− 1323, p<0.01) deteriorated (Figure 5A-D). Consistent with identified decline in established measures, remote monitored mean pulmonary artery pressure (3.7, 1.8-5.5 mmHg, p<0.001), total pulmonary resistance (−1.7, −0.8 − −2.7 WU, p<0.01) and night heart rate (3.5, 0.3-6.6 bpm, p<0.05) were increased and cardiac output (−0.4, −0.2 − −0.7 L/min, p<0.01) and physical activity (−49.8, −46.9 − −65.0 min, p<0.001) decreased (Figure 5E-J). Daily measures of these parameters taken from the patients home demonstrate that mean pulmonary artery pressure and total pulmonary resistance as well as a reduction in cardiac output and physical activity at least ten days prior to a clinical worsening event. In 8 patients changes were apparent at least 15-days prior to a clinical worsening event.

**Figure 5:**
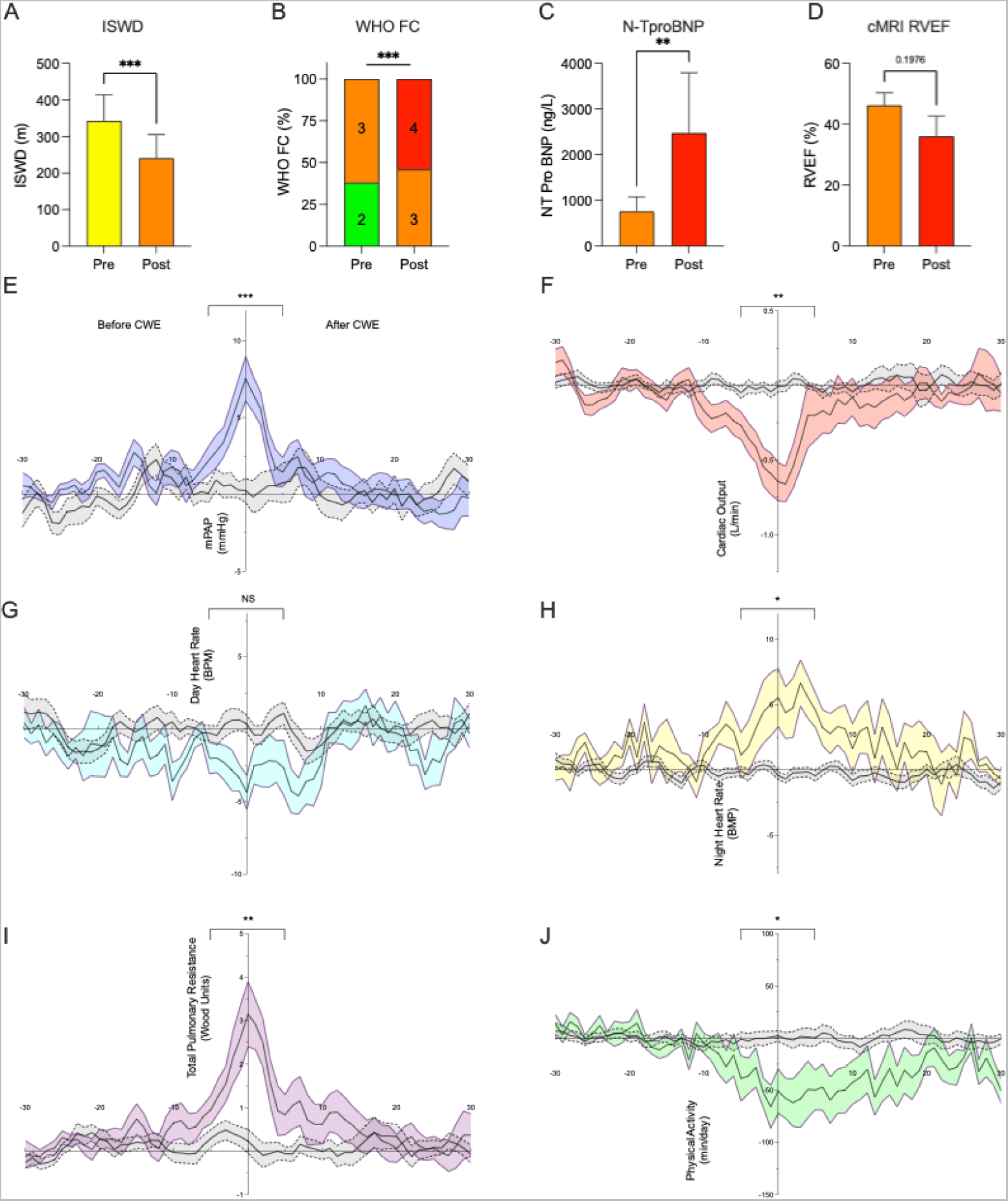
Established risk measures and remote-monitored physiology at the time of a clinically worsening event. A. Incremental shuttle walk distance (ISWD), B. WHO functional class (WHO FC), C. NTProBNP and D. right ventricular ejection fraction from cardiac MRI (cMRI-RVEF). Mean +/−SEM, Wilcoxon, **p<0.01, ***p<0.001 A-C: n=13, D: n=5). E: mean pulmonary artery pressure, F: cardiac output, G: day heart rate, H: night heart rate; I total pulmonary resistance; J: physical activity. Data is presented with treatment change at day 0 with days –30 to day −1 as days preceding, and days +1 to day +30 as days following treatment change. Control group comprises matched 60-day periods without CWE (grey). CWE n=13, mean +/−SEM, *p<0.05, **p<0.01, ***p<0.001 paired Student’s t-test.

### Remote physiological risk score: early evaluation of clinical efficacy and detection of worsening

To establish a summary measure, physiological parameters that change with initiation of disease-specific therapy and clinical worsening were combined to provide a remote physiological risk score. Following a clinically indicated increase in therapy, an improvement in physiology was identifiable at day three using a risk score of parameters from the insertable cardiac monitor and pulmonary artery pressure monitor compared with day 18 using parameters from the insertable cardiac monitor only. Preceding a clinical worsening event, adverse physiology was identifiable at day – 16 using a risk score of parameters from the insertable cardiac monitor and pulmonary artery pressure monitor compared with day −4 using parameters from the insertable cardiac monitor only (Figure 6).

**Figure 6:**
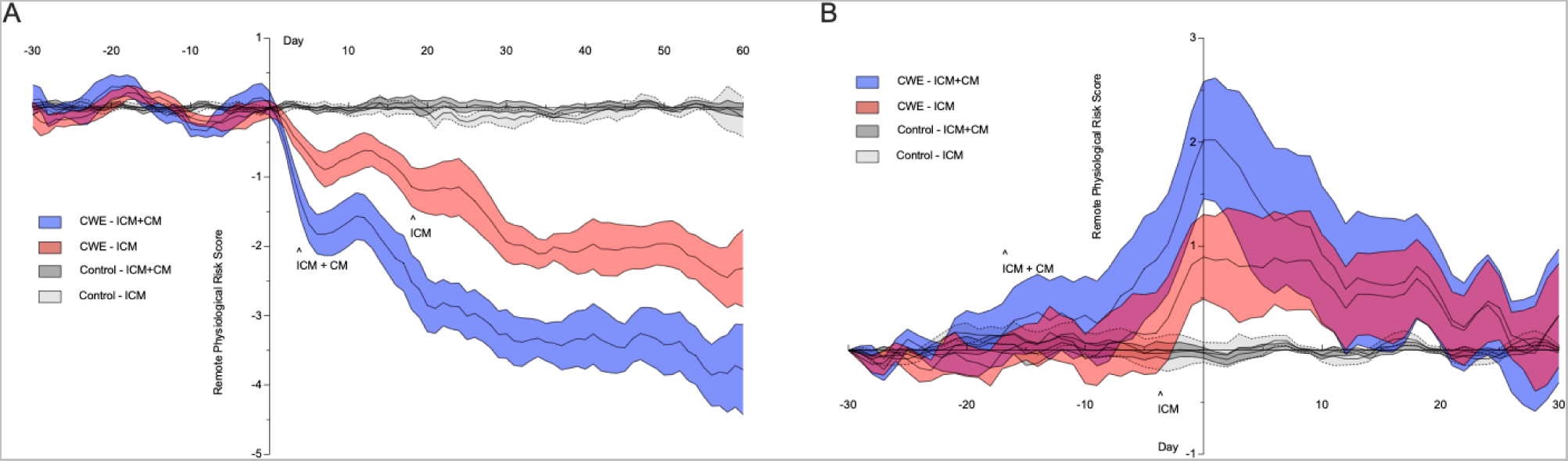
Remote risk score at the time of clinically indicated therapeutic escalation and clinical worsening events. Therapeutic escalation (A) and clinical worsening event (B) occur at day 0 with days –30 to day −1 as days preceding, and days +1 to day +30 or +60 as days following. Control group comprises matched time periods with no TE or CWE (grey). TE n=18, CWE n=13, mean +/−SEM, two-way ANOVA with Dunnett’s correction, ^ indicates earlies point of statistical significance.

### Physiological response to treatment initiation and transition in a patient with PAH

As a demonstration of potential utility, we applied the remote risk score to a single patient’s data through the initiation of disease specific therapy and the transition from intravenous prostanoid to oral therapy. A female in her early 30’s was admitted to hospital following a 12-month period of increasing shortness of breath, reducing exercise tolerance and the recent development of pre-syncope. Following baseline catheterisation a diagnosis of pulmonary arterial hypertension is confirmed and devices implanted, and treatment initiated with sildenafil, ambrisentan and intravenous epoprostenol. TPR and physical activity improve over a one-year period with initiation and up-titration of therapy (Figure 7A). Following 14-months on IV therapy a patient led-decision is made to wean (day 410) and withdraw (day 580) epoprostenol and initiate triple oral therapy with selexipag (day 585, Figure 7B). During the initial dose reduction from 16 ng/kg/min of epoprostenol there is no change in remote measured physiology. Following an inpatient admission with reduction to 9 ng/kg/min there is a dose-dependent worsening in remote measured physiology and remote risk score with down-titration of epoprostenol. With initiation of selexipag there is a dose-dependent improvement in remote measured physiology and remote risk score.

**Figure 7:**
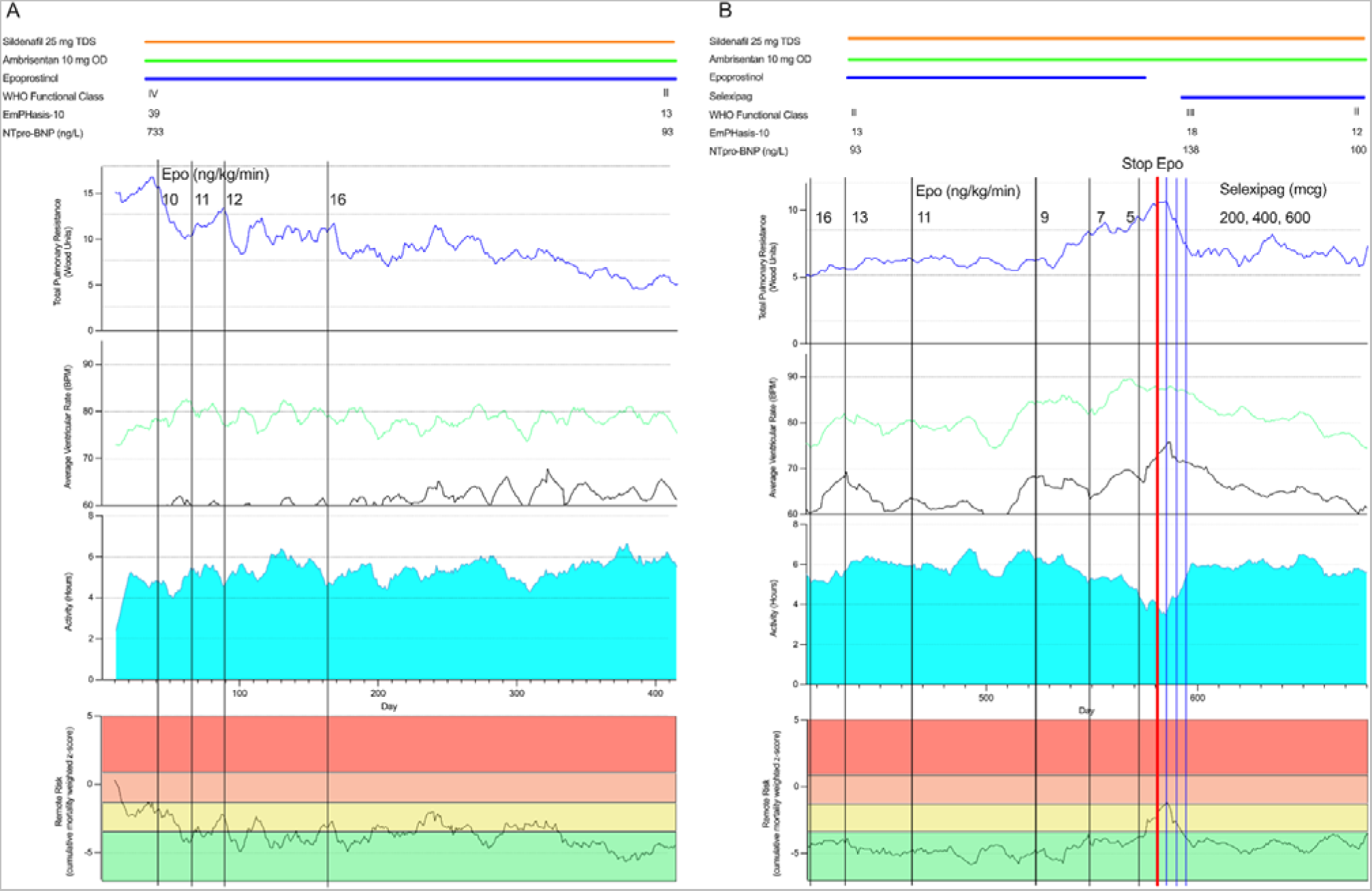
remote monitored physiology in clinical practise. A female in her early 30’s is admitted to hospital following a 12-month period of increasing shortness of breath, reducing exercise tolerance and the recent development of pre-syncope. A: Following a diagnosis of PAH sildenafil, ambrisentan and epoprostenol are started. TPR and physical activity improve gradually over a one-year period. B: A patient led-decision is made to wean (day 410) and withdraw (day 580) epoprostenol and initiate selexipag (da**y** 585). Dose-dependent changes are observed in remote measured physiology and risk with down-titration of epoprostinol and initiation of selexipag. (TPR – blue; night heart rate – black; day heart rate −green; physical activity – light blue; remote risk – bottom panel).

## Discussion

Current guidelines for the diagnosis and management of patients with pulmonary arterial hypertension recommend hospital-based risk evaluation every 3-6 months to facilitate therapeutic optomisation.^11^ Despite this, a low risk profile is not achieved in most patients.^28,29^ In studies investigating the efficacy of novel therapeutics hospital-based invasive and non-invasive testing are undertaken at baseline and after 3-6 months of drug exposure. ^11^ As such, an early, remote means for the evaluation of therapeutic efficacy has the potential to facilitate therapeutic optimisation and personalised therapy and improve clinical study design.

In patients with heart failure (not related to pulmonary arterial hypertension), implanted devices that provide daily, remote measures of cardiovascular physiology may be used for assessment of risk, therapeutic optimisation, and service prioritisation.^14^ To establish a means for remote, early, identification of clinical efficacy and/or clinical worsening, patients with pulmonary arterial hypertension were implanted with two devices to collect remote physiological data. Constant with studies of approved therapies and changes in routine clinical evaluations (ISWD, NT-proBNP, WHO functional class and right ventricular ejection fraction), following a clinically-indicated increase in therapy, a reduction in mean pulmonary artery pressure and total pulmonary resistance was observed at day 7 and 14, followed by increases in cardiac output and physical activity at day 22, and 42. This sequence may reflect a reduction in resistance as a consequence of pulmonary arterial vasodilatation followed by an improvement in cardiac function, that in turn permits increased physical activity. Of particular interest, all parameters were altered in advance of the recommended 3-6 month follow-up time period used in clinical practise and in clinical studies.^11^ Additionally, clinical worsening events were preceded by altered physiology by 16 days, potentially allowing for more timely intervention in deteriorating patients.

Patients with pulmonary hypertension are managed in referral centres often requiring significant travel for evaluation and follow-up. Remote evaluation offers the opportunity to prioritise services, directing resources to patients requiring increased therapy. In patients with heart failure summary scores of physiological parameters measured by implanted devices facilitate assessment of risk, therapeutic optimisation, and service prioritisation.^14^ To form a single summary measure of physiological indicators appropriate for the evaluation of patients with PH the z-score for total pulmonary resistance, heart rate reserve and physical activity were summed. Consistent with the performance of the individual components, a risk score of physiological parameters measured by the insertable cardiac monitor detected change 18 days following an increase in therapy and four days before a clinical worsening event. The risk score of physiological parameters measured by both the insertable cardiac monitor and pulmonary artery pressure monitor improved detection to three days after therapy increase and 18 days before a clinical worsening event. The capacity to rapidly evaluate the effect of treatment decisions offers to impact on early phase clinical trial designs and to change the current care paradigm.

The specific parameters measured in the study require implantation of minimally invasive devices, however, a more limited data set may be monitored using wrist worn or non-invasive technology. Both approaches have benefits and limitations: the implantation of minimally invasive devices is associated with a limited risk^30^ and cost, however provides a secure reliable data collection and transfer system; wrist worn technology can provide an assessment of heart rate, heart rate variability, physical activity and derived measures, however, data collection is not secure and there are limitations with accurate heart rate/variability detection exacerbated by motion artifact, skin pigmentation and poor peripheral perfusion.^31^ The most appropriate technology for the monitoring of patients with PH remains to be determined, however, a hierarchy of non-invasive, minimally invasive, and invasive technology may be appropriate when also considering patient preference, technological availability and access, and disease severity.

In clinical research, the increased frequency of haemodynamic and physical capacity evaluation may permit study designs that are currently limited by the requirement for invasive/hospital-based investigations and provide valuable insight into individualised patient response, dose-response and time-to-effect.^32,33^ This approach is being actively investigated in studies of approved^34^ and repurposed therapies.^35^ Currently approved therapies for the treatment of pulmonary arterial hypertension target vasodilatation and vasoconstriction, however, novel therapies that target the vascular remodelling that drives disease are in development.^36^ It may therefore be possible to demonstrate differential mechanism of action between approved and novel therapies by selective withdrawal of both drug types in individual patients.

The capacity to evaluate the efficacy of a therapy remotely, in a time frame shorter than the recommended 3-6 months offers the opportunity for a personalised approach to therapy balancing therapeutic efficacy against other important factors such as side effects and quality-of-life and to structure clinical studies in a way that provide increased information on dose-response and/or compares therapies head-to-head. Such an approach would reliably, remotely evaluate how a patient feels, functions and survives in real time. Importantly patients found the approach described acceptable and felt that care and evaluation were improved through the use of remote monitoring (PPIE summary).

## Limitations

Changes in remote monitored data were observed preceding and following clinically indicated increases in therapy and clinical worsening events. There was no placebo group and the central Sheffield research team were not blinded. Remote-monitored physiological data from implanted devices was used to provide matched control data in a time-stratified bidirectional case–crossover study providing a study approach that is statistically powerful and efficient in terms of the burden placed on patients. Access to hospital services for scheduled visits was limited due to public health measures put in place to limit the effect of COVID-19 meaning that follow up intervals differed between patients and not all patients with therapeutic escalation clinical worsening events underwent imaging assessment of right ventricular function.

## Conclusions

In patients implanted with an insertable cardiac and pulmonary artery pressure monitor change in physiology was observed following clinically indicated therapeutic escalation and prior to clinical worsening. Remote haemodynamic and cardiac monitoring may facilitate personalised, proactive medicine and innovative clinical study designs.

## Author contributions

JTM and AMKR conceived and designed the study. JTM, SB, HZ, JP, CA, JT, DNN, CB, CP, CR, SR, JA, FV, NH, IA, JH, AH, AC, AJS, AART, RC, CA, DGK, MT and AMKR collected the data. JTM, SB, HZ, JP, CA, JT, DNN, CB, FV, AJS, TB, SKWH, JMSW, AART, RC, CE, DGK and AMKR performed data analysis and interpretation. All authors provided substantial input to the manuscript and approved the final version.

## Data Availability

All data produced in the present study are available upon reasonable request to the authors

---

**Supplemental Table 1:** Serious adverse events and adverse events reported in the FIT-PH study.

| Adverse Events | Number of occurrences | Action taken / further information |
| --- | --- | --- |
| Serious Adverse Events | 0 | NA |
| Bleeding | 1 | Minor haemoptysis, resolved with no intervention |
| Device erosion | 1 | Explanted, allowed to heal and new device implanted |
| Infection | 0 | NA |
| Vascular injury | 0 | NA |
| Pulmonary Embolism | 0 | NA |
| Drug interaction | 1 | Selexipag and clopidogrel.<br>Clopidogrel replaced with prasugrel. |

**Supplemental Figure 1:**
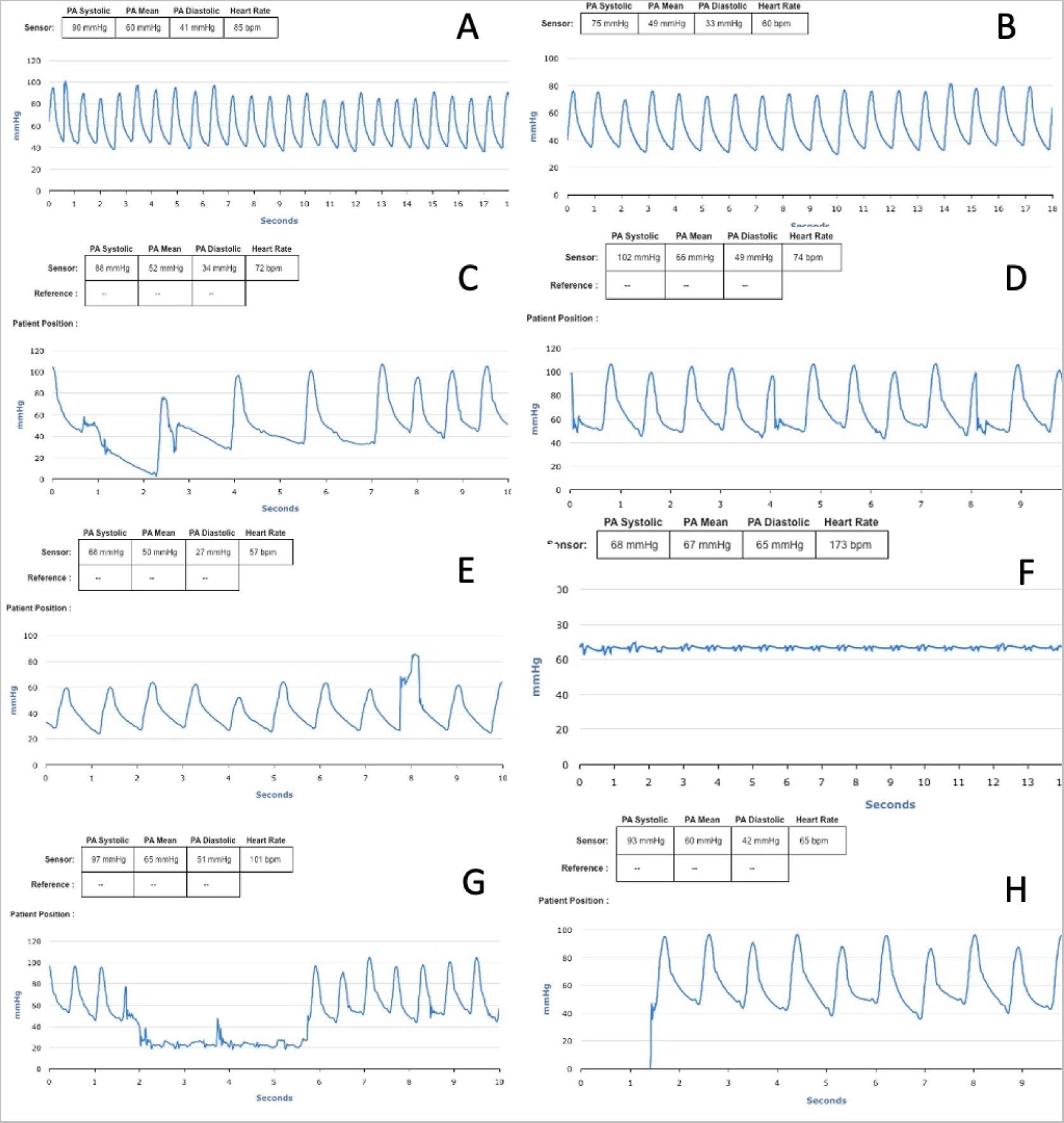
Examples of non-clinically significant ‘suspect’ pulmonary artery pressure waveforms measured from a CardioMEMS device resulting from: A: Non-rested physiological state; B: rested physiological state in the same patient; C: a pause followed by a compensatory bradycardia; D: Frequent ventricular ectopy; E: Non physiological waveform; F: Incorrect waveform frequency detected; G: Waveform damping; H: Non transmission of waveforms.

**Supplemental Figure 2:**
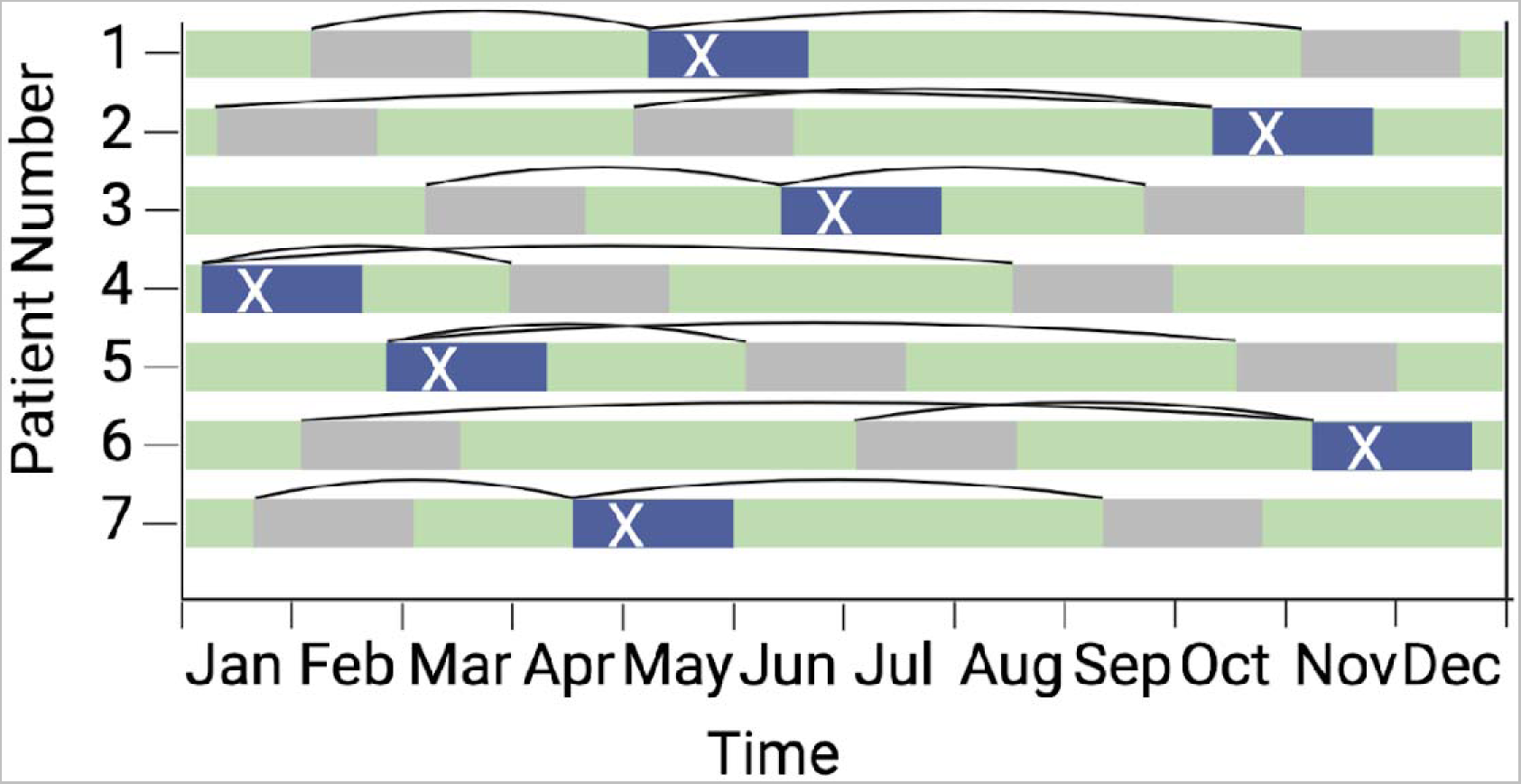
Representative sampling of remote monitoring data for event and control periods in time-stratified bidirectional case–crossover study. Period of remote monitoring is indicated by green. Timepoints of clinician-directed therapeutic escalation or clinical worsening events are indicated by white X with the time 30-days preceding and 30-days (CWE) or 60-days (TE) following the event indicated in blue. Control time periods are indicated in grey.

**Supplemental Figure 3:**
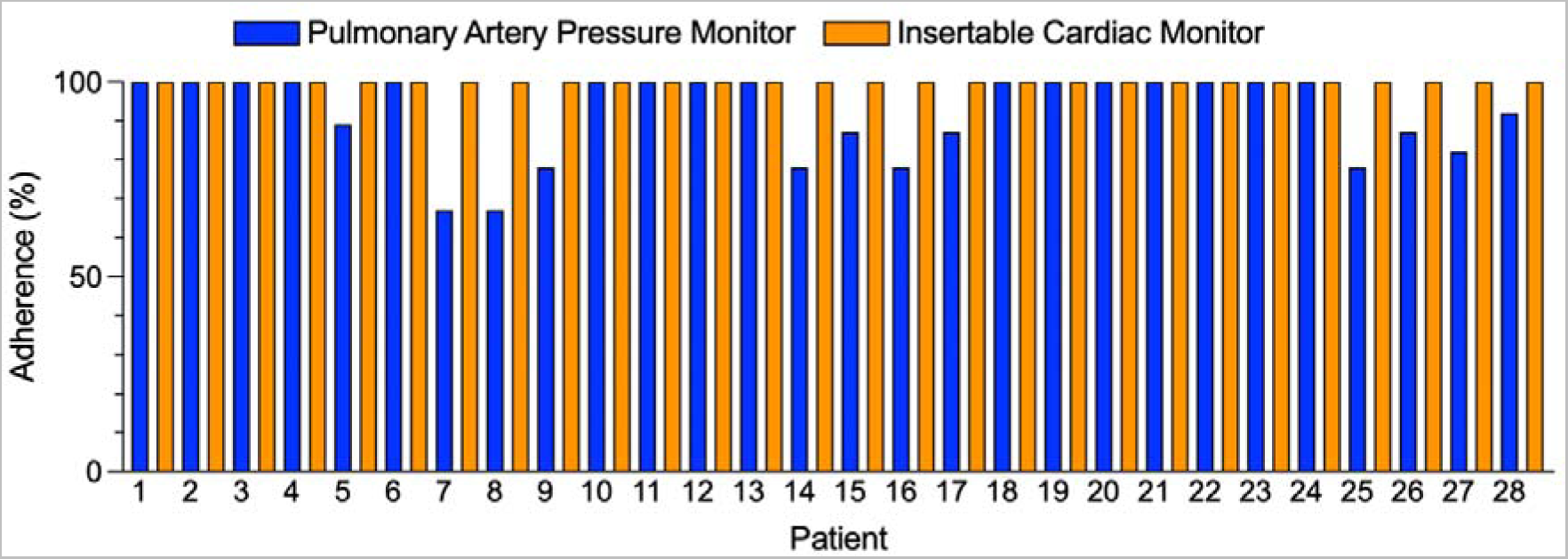
Reading adherence in patients with insertable cardiac and pulmonary artery pressure monitors. Percentage of weekly readings completed by individual patients over the first three months following implantation for insertable cardiac monitor (orange) and pulmonary artery pressure monitor (blue).

## References

1. PHA UK. What it means to live with PH today. https://www.phauk.org/research-survey-work/living-ph-report/.

2. PHA UK. Helping people get the most from their PH treatments: Phoenix study proposal findings. https://www.phauk.org/app/uploads/2021/04/Phoenix-study-research-report.pdf.

3. Kiely DG, Elliot CA, Sabroe I, Condliffe R. Pulmonary hypertension: diagnosis and management. Br Med J 2013:346–357.

4. Humbert M, Sitbon O, Simonneau G. Treatment of Pulmonary Arterial Hypertension. New England Journal of Medicine 2004;351:1425–1436.

5. Thenappan T, Shah SJ, Rich S, Tian L, Archer SL, Gomberg-Maitland M. Survival in pulmonary arterial hypertension: a reappraisal of the NIH risk stratification equation. Eur Respir J 2010;35:1079–1087.

6. Ling Y, Johnson MK, Kiely DG, Condliffe R, Elliot CA, Gibbs JSR, Howard LS, Pepke-Zaba J, Sheares KKK, Corris PA, Fisher AJ, Lordan JL, Gaine S, Coghlan JG, Wort SJ, Gatzoulis MA, Peacock AJ. Changing demographics, epidemiology, and survival of incident pulmonary arterial hypertension: Results from the pulmonary hypertension registry of the United Kingdom and Ireland. Am J Respir Crit Care Med 2012;186:790–796.

7. Benza RL, Miller DP, Barst RJ, Badesch DB, Frost AE, McGoon MD. An Evaluation of Long-term Survival From Time of Diagnosis in Pulmonary Arterial Hypertension From the REVEAL Registry. Chest 2012;142:448–456.

8. Halliday SJ, Hemnes AR. Identifying “super responders” in pulmonary arterial hypertension. Pulm Circ 2017;7:300–311.

9. Pulido T, Adzerikho I, Channick RN, Delcroix M, Galiè N, Ghofrani H-A, Jansa P, Jing Z-C, Brun F-O Le, Mehta S, Mittelholzer CM, Perchenet L, Sastry BKS, Sitbon O, Souza R, Torbicki A, Zeng X, Rubin LJ, Simonneau G. Macitentan and Morbidity and Mortality in Pulmonary Arterial Hypertension. New England Journal of Medicine 2013;369:809–818.

10. Sitbon O, Channick RN, Chin KM, Frey A, Gaine S, Galie N, Ghofrani H-A, Hoeper MM, Lang IM, Preiss R, Rubin LJ, Scala L Di, Tapson V, Adzerikho I, Liu J, Moiseeva O, Zeng X, Simonneau G, McLaughlin V V. Selexipag for the treatment of pulmonary arterial hypertension. New England Journal of Medicine 2015;373:2522–2533.

11. Humbert M, Kovacs G, Hoeper MM, Badagliacca R, Berger RMF, Carlsen J, Mayer E, Nagavci B, Olsson KM, Pepke-zaba J, Quint JK, Simonneau G, Sitbon O, Toshner M. 2022 ESC / ERS Guidelines for the diagnosis and treatment of pulmonary hypertension Developed by the task force for the diagnosis and treatment of (ESC) and the European Respiratory Society (ERS). Endorsed by the International Society for Heart and Lu. Eur Heart J 2022:1–114.

12. Hoeper MM, Pausch C, Olsson KM, Huscher D, Pittrow D, Grünig E, Staehler G, Vizza CD, Gall H, Distler O, Opitz C, Gibbs JSR, Delcroix M, Ghofrani HA, Park D-H, Ewert R, Kaemmerer H, Kabitz H-J, Skowasch D, Behr J, Milger K, Halank M, Wilkens H, Seyfarth H-J, Held M, Dumitrescu D, Tsangaris I, Vonk-Noordegraaf A, Ulrich S, Klose H, Claussen M, Lange TJ, Rosenkranz S. COMPERA 2.0: A refined 4-strata risk assessment model for pulmonary arterial hypertension. European Respiratory Journal 2021:2102311.

13. Weatherald J, Boucly A, Sahay S, Humbert M, Sitbon O. The low-risk profile in pulmonary arterial hypertension: Time for a paradigm shift to goal-oriented clinical trial endpoints? Am J Respir Crit Care Med 2018;197:860–868.

14. Lindenfeld J, Zile MR, Desai AS, Bhatt K, Ducharme A, Horstmanshof D, Krim SR, Maisel A, Mehra MR, Paul S, Sears SF, Sauer AJ, Smart F, Zughaib M, Castaneda P, Kelly J, Johnson N, Sood P, Ginn G, Henderson J, Adamson PB, Costanzo MR. Haemodynamic-guided management of heart failure (GUIDE-HF): a randomised controlled trial. The Lancet 2021;398:991–1001.

15. Brugts JJ, Radhoe SP, Clephas PRD, Aydin D, Gent MWF van, Szymanski MK, Rienstra M, Heuvel MH van den, Fonseca CA da, Linssen GCM, Borleffs CJW, Boersma E, Asselbergs FW, Mosterd A, Brunner-La Rocca H-P, Boer RA de, Emans ME, Beeres SLMA, Heerebeek L, Kirchhof C, Ramshorst J Van, Spee R, Smilde T, Eck M Van, Kaplan E, Hazeleger R, Tukkie R, Feenema M, Kok W, Halm V Van, Handoko ML, Kimmenade R Van, Post M, Mieghem N Van, Manintveld OC. Remote haemodynamic monitoring of pulmonary artery pressures in patients with chronic heart failure (MONITOR-HF): a randomised clinical trial. The Lancet 2023;6736:1–11.

16. Sanna T, Diener H-C, Passman RS, Lazzaro V Di, Bernstein RA, Morillo CA, Rymer MM, Thijs V, Rogers T, Beckers F, Lindborg K, Brachmann J. Cryptogenic Stroke and Underlying Atrial Fibrillation. New England Journal of Medicine 2014.

17. Bonnesen MP, Frodi DM, Haugan KJ, Kronborg C, Graff C, Højberg S, Køber L, Krieger D, Brandes A, Svendsen JH, Diederichsen SZ. Day-to-day measurement of physical activity and risk of atrial fibrillation. Eur Heart J 2021;42:3979–3988.

18. Tree M, White J, Midha P, Kiblinger S, Yoganathan A. Validation of cardiac output as reported by a permanently implanted wireless sensor. Journal of Medical Devices, Transactions of the ASME 2016;10:1–7.

19. Karamanoglu M, Bennett T, Ståhlberg M, Splett V, Kjellström B, Linde C, Braunschweig F. Estimation of cardiac output in patients with congestive heart failure by analysis of right ventricular pressure waveforms. Biomed Eng Online 2011;10:1–13.

20. Benza RL, Doyle M, Lasorda D, Parikh KS, Correa-Jaque P, Badie N, Ginn G, Airhart S, Franco V, Kanwar MK, Murali S, Raina A, Agarwal R, Rajagopal S, White J, Biederman R. Monitoring Pulmonary Arterial Hypertension Using an Implantable Hemodynamic Sensor. Chest 2019;156:1176–1186.

21. Conraads VM, Spruit MA, Braunschweig F, Cowie MR, Tavazzi L, Borggrefe M, Hill MRS, Jacobs S, Gerritse B, Veldhuisen DJ Van. Physical activity measured with implanted devices predicts patient outcome in chronic heart failure. Circ Heart Fail 2014;7:279–287.

22. Cowie MR, Sarkar S, Koehler J, Whellan DJ, Crossley GH, Tang WHW, Abraham WT, Sharma V, Santini M. Development and validation of an integrated diagnostic algorithm derived from parameters monitored in implantable devices for identifying patients at risk for heart failure hospitalization in an ambulatory setting. Eur Heart J 2013;34:2472–2480.

23. Shoemaker MJ, Cartwright K, Hanson K, Serba D, Dickinson MG, Kowalk A. Concurrent Validity of Daily Activity Data from Medtronic ICD/CRT Devices and the Actigraph GT3X Triaxial Accelerometer: A Pilot Study. Cardiopulm Phys Ther J 2017;28:3–11.

24. Pressler A, Danner M, Esefeld K, Haller B, Scherr J, Schömig A, Halle M, Kolb C. Validity of cardiac implantable electronic devices in assessing daily physical activity. Int J Cardiol 2013;168:1127–1130.

25. Galiè N, Barberà JA, Frost AE, Ghofrani H-A, Hoeper MM, McLaughlin V V., Peacock AJ, Simonneau G, Vachiery J-L, Grünig E, Oudiz RJ, Vonk-Noordegraaf A, White RJ, Blair C, Gillies H, Miller KL, Harris JHN, Langley J, Rubin LJ. Initial Use of Ambrisentan plus Tadalafil in Pulmonary Arterial Hypertension. New England Journal of Medicine 2015;373:834–844.

26. Mostofsky E, Coull BA, Mittleman MA. Analysis of Observational Self-matched Data to Examine Acute Triggers of Outcome Events with Abrupt Onset. 2018;29.

27. Sitbon O, Benza RL, Badesch DB, Barst RJ, Elliott CG, Gressin V, Lemarié JC, Miller DP, Rouzic EM Le, Simonneau G, Frost AE, Farber HW, Humbert M, McGoon MD. Validation of two predictive models for survival in pulmonary arterial hypertension. European Respiratory Journal 2015;46:152–164.

28. Boucly A, Weatherald J, Savale L, Jaïs X, Cottin V, Prevot G, Picard F, Groote P De, Jevnikar M, Bergot E, Chaouat A, Chabanne C, Bourdin A, Parent F, Montani D, Simonneau G, Humbert M, Sitbon O. Risk assessment, prognosis and guideline implementation in pulmonary arterial hypertension. European Respiratory Journal 2017;50:1–10.

29. Hoeper MM, Kramer T, Pan Z, Eichstaedt CA, Spiesshoefer J, Benjamin N, Olsson KM, Meyer K, Vizza CD, Vonk-Noordegraaf A, Distler O, Opitz C, Gibbs JSR, Delcroix M, Ghofrani HA, Huscher D, Pittrow D, Rosenkranz S, Grünig E. Mortality in pulmonary arterial hypertension: prediction by the 2015 European pulmonary hypertension guidelines risk stratification model. European Respiratory Journal 2017;50:1700740.

30. Lin AL, Hu G, Dhruva SS, Kinard M, Redberg RF. Quantification of Device-Related Event Reports Associated with the CardioMEMS Heart Failure System. Circ Cardiovasc Qual Outcomes 2022;15:E009116.

31. Cabanas AM, Fuentes-Guajardo M, Latorre K, León D, Martín-Escudero P. Skin Pigmentation Influence on Pulse Oximetry Accuracy: A Systematic Review and Bibliometric Analysis. Sensors (Basel) 2022;22.

32. Kane PB, Bittlinger M, Kimmelman J. Individualized therapy trials: navigating patient care, research goals and ethics. Nat Med 2021;27:1679–1686.

33. Senn S. Statistical pitfalls of personalized medicine. Nature 2018;563:619–621.

34. NIH. Pulmonary Hypertension: Intensification and Personalisation of Combination Rx (PHoenix). *Clinicaltrials.gov*. https://classic.clinicaltrials.gov/ct2/show/NCT05825417 (26 June 2023)

35. Wilkins MR, Mckie MA, Law M, Roussakis AA, Harbaum L, Church C, Coghlan JG, Condliffe R, Howard LS, Kiely DG. Positioning imatinib for pulmonary arterial hypertension: A phase I/II design comprising dose finding and single-arm efficacy. Pulm Circ 2021;11:20458940211052824.

36. Hoeper MM, Badesch DB, Ghofrani HA, Gibbs JSR, Gomberg-Maitland M, McLaughlin V V, Preston IR, Souza R, Waxman AB, Grünig E, Kopeć G, Meyer G, Olsson KM, Rosenkranz S, Xu Y, Miller B, Fowler M, Butler J, Koglin J, Oliveira Pena J de, Humbert M. Phase 3 Trial of Sotatercept for Treatment of Pulmonary Arterial Hypertension. N Engl J Med 2023;388:1478–1490.

